# Narcolepsy is associated with cardiovascular burden

**DOI:** 10.64898/2026.04.22.26351468

**Authors:** Reyhane Eghtedarian, Hele Haapaniemi, Markus Ramste, FinnGen, Hanna M. Ollila

## Abstract

**Background:** Narcolepsy is a debilitating sleep disorder caused by hypocretin deficiency. Aside from its role to induce wakefulness, hypocretin is linked to modulated appetite and metabolism, often resulting in weight gain.

**Study objectives:** We aimed to unravel the comprehensive epidemiological connection between narcolepsy and major cardiometabolic outcomes.

**Methods:** We analyzed cardiovascular and metabolic disease distribution in the FinnGen study. Using longitudinal electronic health records, we assessed associations between narcolepsy, cardiac/metabolic markers, and prescriptions for relevant drugs.

**Results:** Our findings demonstrate significant associations between narcolepsy and metabolic traits (OR [95% CI] = 2.65 [1.81, 3.89]) as well as stroke (OR = 2.36 [1.38, 4.04]). Narcolepsy patients exhibit a less favourable metabolic profile, including higher glucose levels (OR = 1.1143 [1.0599, 1.1715]) and dyslipidaemia. This is supported by increased prescriptions of insulin (OR = 2.269 [1.46, 3.53]), simvastatin (OR = 2.292 [1.59, 3.31]), and metformin (OR = 2.327 [1.66, 3.25]), reflecting high metabolic disturbances. Furthermore, positive associations with antihypertensive and antiplatelet medications were observed, consistent with elevated cardiovascular risk.

**Conclusion:** Taken together, our findings highlight the cardiometabolic burden in narcolepsy. This study enhances understanding of the metabolic and cardiovascular consequences of narcolepsy and offers timely guidance for effective disease control.

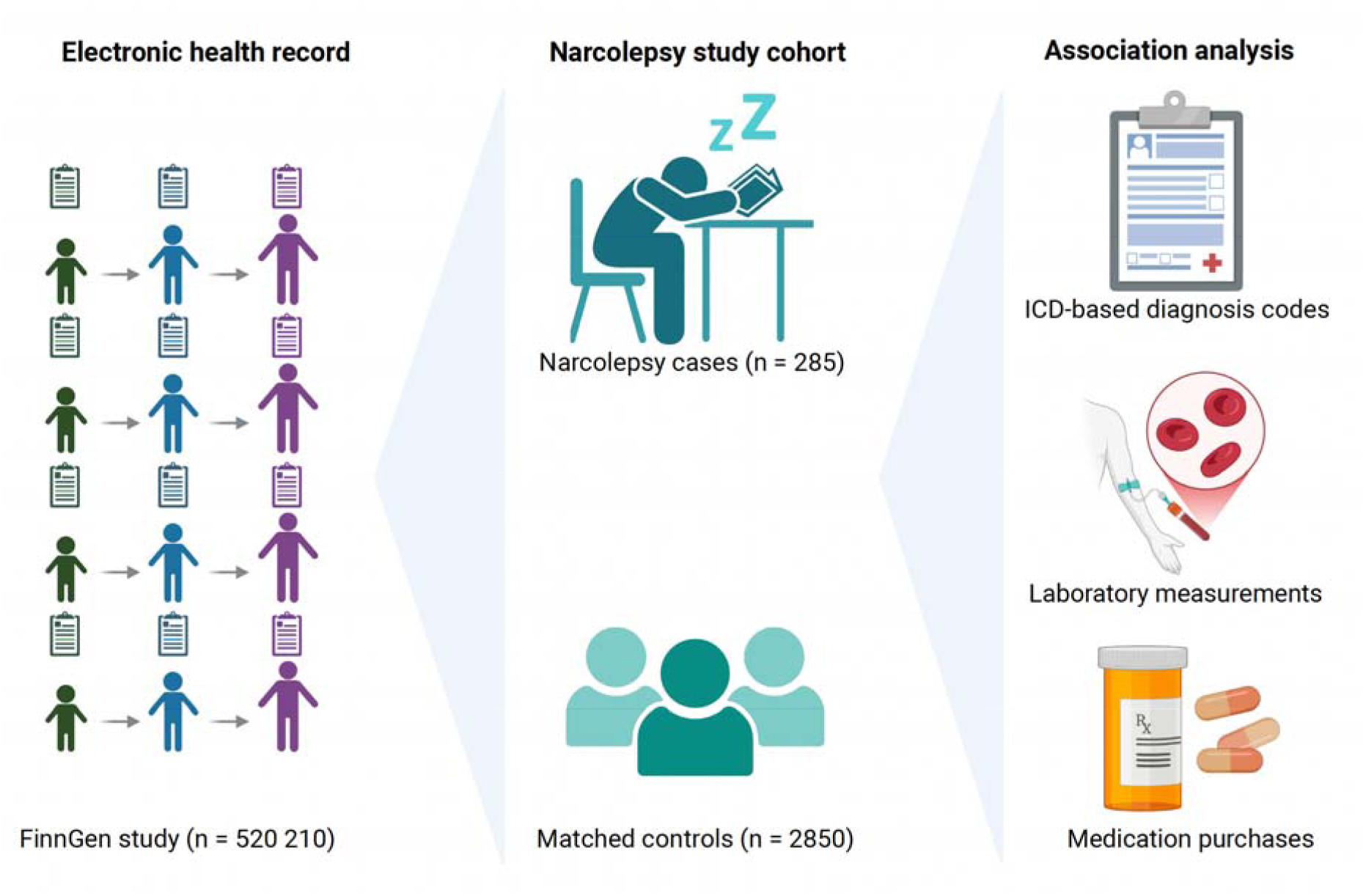

## Introduction

Narcolepsy is a chronic, neurological sleep disorder characterized by excessive daytime sleepiness, sleep fragmentation, sleep paralysis, and cataplexy (a sudden loss of muscle tone, triggered by strong, often positive emotions) ^1,2^. The first symptoms typically occur between 10 and 25 years of age^3^. According to the definition of International classification of sleep disorders (ICSD-3), narcolepsy is classified in type 1 (NT1) and type 2 (NT2). NT1 is characterized by cataplexy and/or lack of hypocretin 1 in their cerebrospinal fluid, whereas NT2 is defined by the absence of cataplexy and normal hypocretin 1 level^4^. The global prevalence of narcolepsy is estimated to be 25-50 per 100,000 individuals^5–9^.

While hypocretins are primarily recognized to regulate sleep-wake transition^10^, they have been implicated to affect appetite regulation and energy balance^11,12^, control autonomic functions such as heart rate (HR) and blood pressure (BP), and modulate neuroendocrine system ^13,14^. These diverse roles help explain metabolic disorders including obesity, diabetes, and cardiovascular diseases (CVDs), often observed in narcolepsy.

CVDs are a group of non-communicable diseases, which are attributed to the heart and the vasculature system^15^, including coronary heart disease (CHD), coronary artery disease (CAD), peripheral artery disease (PAD), stroke, aortic aneurysm, atherosclerosis, and heart failure. CVDs are the leading causes of mortality and morbidity worldwide, with over 20 million deaths in 2021, accountable for one third of the global deaths^16^. Well-established risk factors include age, gender, smoking, high blood pressure, high cholesterol and obesity^17,18^. In addition to the traditional risk factors, the effect of sleep on cardiovascular health has gained significant attention. The risk of CVDs is increased by 7%, 26% and 20% in people with snoring, insomnia, and narcolepsy, respectively^19^.

Beyond their established role as risk factors for CVDs, metabolic disorders such as obesity, type 2 diabetes (T2D), and metabolic syndrome represent major global health burdens, with steadily increasing prevalence. The prevalence of these conditions has been reported to be higher among patients with narcolepsy, potentially reflecting the loss of hypocretin neurons involved in the regulation of energy homeostasis^20,21^.

In addition to the direct effects of narcolepsy on cardiometabolic risk, pharmacological treatments used to alleviate symptoms, particularly excessive daytime sleepiness, may further influence cardiac health^22^. For example, modafinil, which is used to promote wakefulness, has been associated with increased antihypertensive medication intake, suggesting a higher prevalence of hypertension among treated patients^22,23^. Similarly, methylphenidate used to treat excessive daytime sleepiness, has been shown to increase heart rate and blood pressure^23^. Conversely, both modafinil and methylphenidate have been reported to reduce appetite, food cravings, and overall food intake, which may lower weight gain and obesity-related risk^24,25^.

In the current work, to further elucidate the link between narcolepsy and cardiometabolic dysfunction, we analyzed electronic health record (EHR) data from the FinnGen study, a cohort encompassing over 500,000 Finnish participants. By integrating longitudinal laboratory measurements and drug prescriptions, we assessed the relationship between narcolepsy and key cardiometabolic biomarkers, and the use of related medications. Our findings aim to refine clinical approaches for comorbidity management, risk stratification, and more personalized preventive and therapeutic interventions.

## Material and methods

### FinnGen narcolepsy cohort

FinnGen, is a large study which includes genotypic and phenotypic data of 520,210 Finns, encompassing EHR, including International Classification of Diseases (ICD) codes (derived from primary care registers, hospital inpatient and outpatient visits), drug prescriptions spanning an individual’s entire lifespan, and laboratory measurement values starting from 2014^26^. Using data from FinnGen release 13 (R13), we identified 285 individuals with ICD10 code G47.4 and ICD9 code 347.1A from outpatients, inpatient registries and primary care, corresponding to narcolepsy and cataplexy.

Based on the FinnGen population, we calculated an incidence rate of 55 narcolepsy cases per 100,000 inhabitants, which corresponds to the general estimates of narcolepsy prevalence in earlier studies^4^. We first performed a descriptive characterization of the cohort to understand the demographic profile, including age, sex and year of diagnosis in narcolepsy patients. Females were predominant in cases (57.5 %) compared to men (38.6 %) (**Figure 1**A). We observed the highest incidence of narcolepsy among people aged 20-30 years, with a median of 37 years (**Figure 1B**). In addition, a variation in annual incidence of narcolepsy was noted, with a peak around 2011 and following years (**Figure 1C**), indicating the known connection between narcolepsy and H1N1 vaccination^27^. To select a subset of controls with similar characteristics to our narcolepsy patients and reduce confounding effects, we matched the cases and the controls based on age and sex, using RStudio 4.5.2 and MatchIt library version 4.7.2, to have a final cohort of 285 cases and 2850 controls.

**Figure 1.**
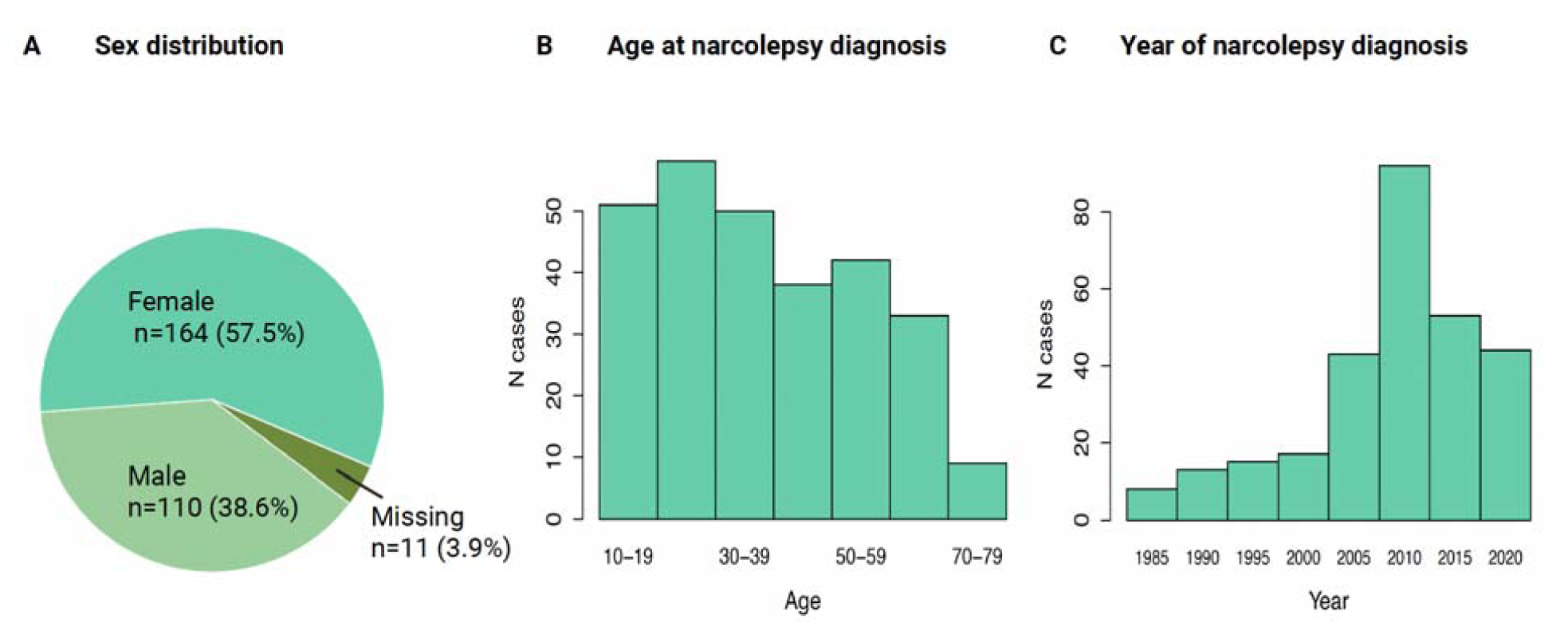
Demographic data for the narcolepsy cohort. A) Distribution of sex within the FinnGen R13 narcolepsy cohort. B) Age at diagnosis of narcolepsy. C) Year of narcolepsy diagnosis.

### FinnGen cardiometabolic cohort

Next, we extracted the cardiometabolic status of the narcolepsy cases and controls, using the ICD9 and ICD10 code diagnoses. For this end, we followed two approaches;

1. First, we used the ICD codes individually corresponding to atherosclerosis, aortic aneurysm, cardiomyopathy, coronary heart disease (CHD), peripheral artery disease (PAD), hypertrophic cardiomyopathy, hypertension, myocardial infarction (MI) or unstable angina, cardiac arrest, T2D, obesity, myocarditis, stroke, heart failure, fatal events including CHD, sudden death of unknown reasons and deaths with myocardial infarction.
2. Second, we grouped the phenotypes into atherosclerosis phenotypes, multi-etiology traits and metabolic traits according to **Table 1**, however, some of the phenotypes remained uncategorized. Conditions having less than five overlapping cases (cardiac arrest and hypertrophic cardiomyopathy) with narcoleptic patients were excluded from the individual phenotype-level analyses to ensure robustness and anonymity of the participants but maintained in the grouped phenotype analyses.

**Table 1.**
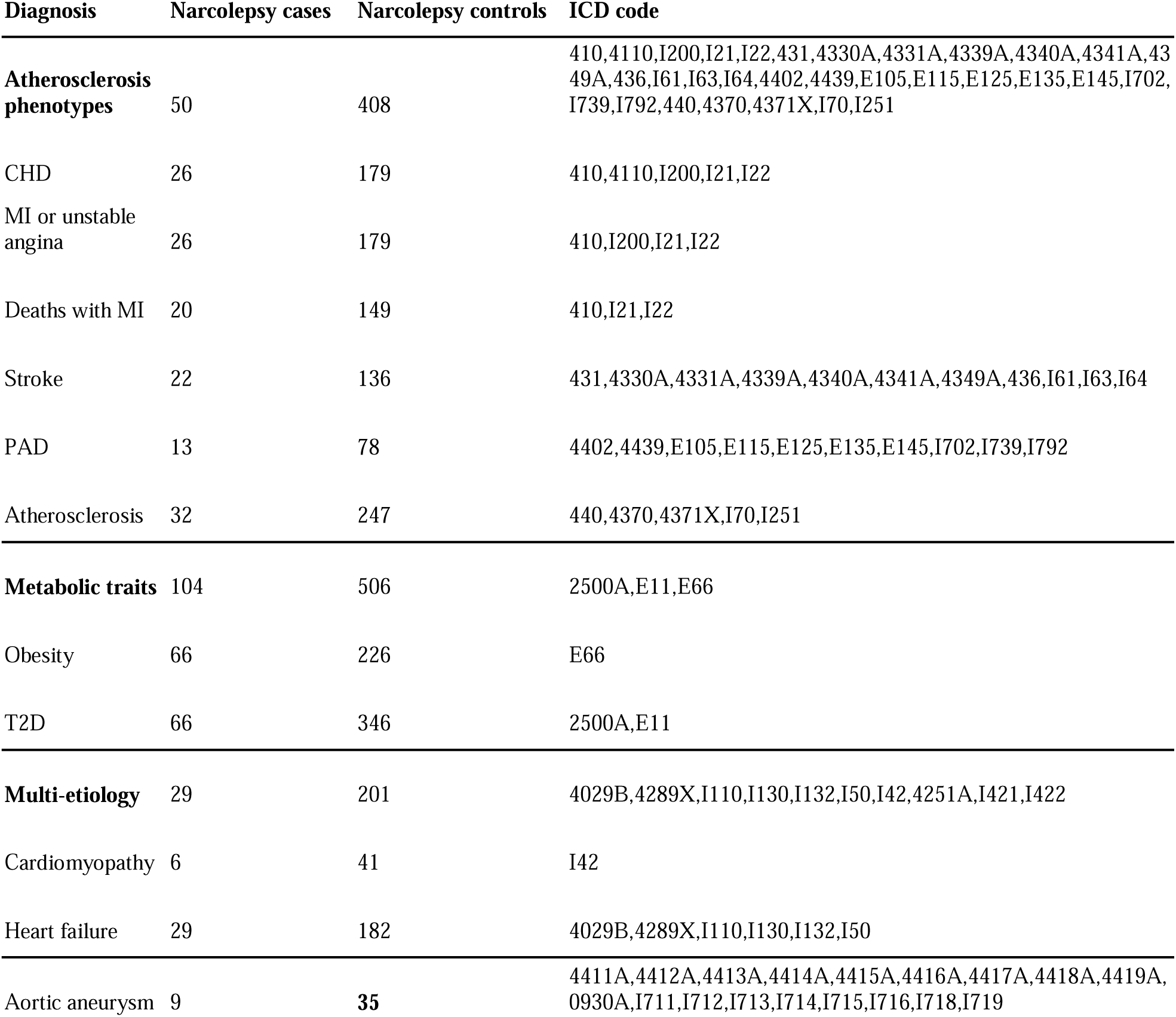
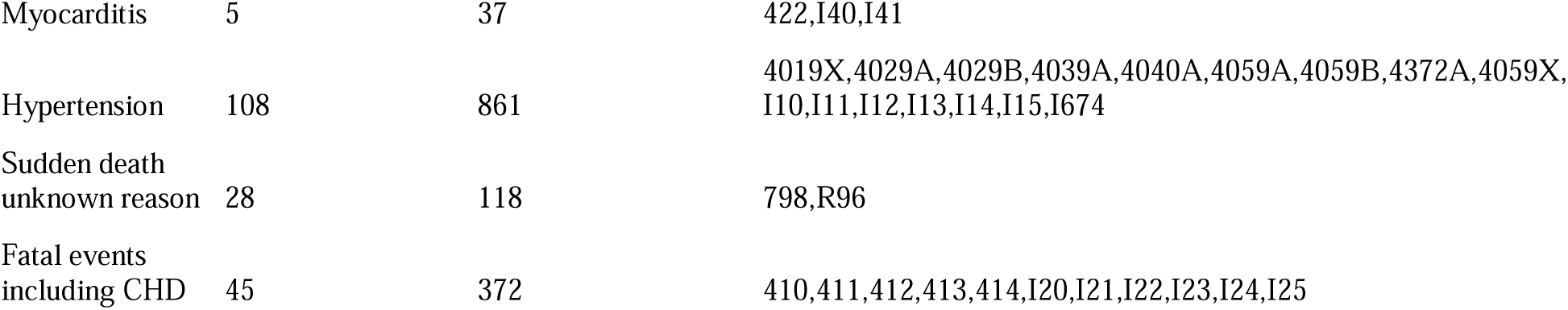
ICD-based codes for the cardiovascular diagnoses.

| Diagnosis | Narcolepsy cases | Narcolepsy controls | ICD code |
| --- | --- | --- | --- |
| **Atherosclerosis phenotypes** | 50 | 408 | 410,4110,I200,I21,I22,431,4330A,4331A,4339A,4340A,4341A,4349A,436,I61,I63,I64,4402,4439,E105,E115,E125,E135,E145,I702,I739,I792,440,4370,4371X,I70,I251 |
| CHD | 26 | 179 | 410,4110,I200,I21,I22 |
| MI or unstable angina | 26 | 179 | 410,I200,I21,I22 |
| Deaths with MI | 20 | 149 | 410,I21,I22 |
| Stroke | 22 | 136 | 431,4330A,4331A,4339A,4340A,4341A,4349A,436,I61,I63,I64 |
| PAD | 13 | 78 | 4402,4439,E105,E115,E125,E135,E145,I702,I739,I792 |
| Atherosclerosis | 32 | 247 | 440,4370,4371X,I70,I251 |
| **Metabolic traits** | 104 | 506 | 2500A,E11,E66 |
| Obesity | 66 | 226 | E66 |
| T2D | 66 | 346 | 2500A,E11 |
| **Multi-etiology** | 29 | 201 | 4029B,4289X,I110,I130,I132,I50,I42,4251A,I421,I422 |
| Cardiomyopathy | 6 | 41 | I42 |
| Heart failure | 29 | 182 | 4029B,4289X,I110,I130,I132,I50 |
| Aortic aneurysm | 9 | **35** | 4411A,4412A,4413A,4414A,4415A,4416A,4417A,4418A,4419A,0930A,I711,I712,I713,I714,I715,I716,I718,I719 |
| Myocarditis | 5 | 37 | 422,I40,I41 |
| Hypertension | 108 | 861 | 4019X,4029A,4029B,4039A,4040A,4059A,4059B,4372A,4059X,I10,I11,I12,I13,I14,I15,I674 |
| Sudden death<br>unknown reason | 28 | 118 | 798,R96 |
| Fatal events<br>including CHD | 45 | 372 | 410,411,412,413,414,I20,I21,I22,I23,I24,I25 |

### Cardiometabolic disorders in narcolepsy

Epidemiological analyses were conducted in the FinnGen computing environment (Sandbox) using RStudio version 4.5.2. Logistic regression was performed using a generalized linear model. Analysis was adjusted with age, sex and ten first genetic principal components (PCs) to adjust for population structure. A separate analysis was performed, including body mass index (BMI) as an additional covariate, to further account for the confounding effect of BMI on cardiometabolic status.

### Kanta laboratory values

Laboratory values were obtained from the Kanta laboratory dataset, released to the FinnGen Sandbox in August 2024. Kanta is a digital service for storing social welfare and healthcare data in the Finnish population (www.kanta.fi). The dataset includes an average of 482 tests taken between 2014 and 2023 per individual for approximately 482,000 living FinnGen participants. Laboratory results are publicly accessible via the LabWAS section of Risteys (https://risteys.finngen.fi/), an online tool to explore phenotype level data from FinnGen.

We matched the unique Observational Medical Outcomes Partnership (OMOP) Common Data Model (CDM) IDs, standardizing healthcare content and terminology^28^, to the test names provided in Kanta lab mapping tool (https://finngen.github.io/kanta_lab_harmonisation_public)/. We selected clinically relevant laboratory values among patients with cardiometabolic disorders, including: fasting glucose [mmol/l], Hemoglobin A1c/Hemoglobin total in blood [mmol/mol], fasting triglycerides (TG) [mmol/l], total cholesterol [mmol/l], HDL cholesterol [mmol/l], LDL cholesterol [mmol/l], Alanine aminotransferase (ALT) enzymatic activity [u (enzymatic activity)/l], Aspartate aminotransferase (AST) enzymatic activity [u/l], Gamma-glutamyl transferase (GGT) enzymatic activity [u/l], erythrocyte sedimentation rate (ESR) Westergren method [mm/h], C-reactive protein (CRP) [mg/l], N-terminal pro-B-type natriuretic peptide (proBNP) [ng/l], creatinine [umol/l], and creatine kinase [u (enzymatic activity)/l], cardiac Troponin T (TnT) [ng/l], cardiac Troponin I (TnI) [ng/l], to study their association with narcolepsy in our cohort. The list of the OMOP IDs and their respective definition of the lab tests, and the number of cases and controls with a valid lab measurement are listed in **Table 2**.

**Table 2.** Lab measurements in narcolepsy cohort.

| Test name | OMOP ID | Narcolepsy cases with lab test | Narcolepsy controls with lab test |
| --- | --- | --- | --- |
| Fasting_glc | OMOP_3018251 | 237 | 1983 |
| HbA1c | OMOP_3004410 | 211 | 1830 |
| Fasting_TG | OMOP_3048773 | 214 | 1972 |
| Cholesterol | OMOP_3019900 | 225 | 2068 |
| Cholesterol HDL | OMOP_3023602 | 222 | 2048 |
| Cholesterol LDL | OMOP_3001308 | 225 | 2087 |
| ALT | OMOP_3006923 | 270 | 2351 |
| AST | OMOP_3013721 | 101 | 706 |
| GGT | OMOP_3026910 | 172 | 1505 |
| ESR | OMOP_3013707 | 199 | 1597 |
| CRP | OMOP_3020460 | 221 | 1858 |
| ProBNP | OMOP_3029187 | 68 | 411 |
| Creatinine | OMOP_3020564 | 261 | 2368 |
| Creatinine kinase | OMOP_3007220 | 93 | 554 |
| Troponin T | OMOP_3019800 | 64 | 399 |
| Troponin I | OMOP_3021337 | 34 | 220 |

### Analysis of laboratory measurements

We investigated the association between selected laboratory values relevant to cardiometabolic health and narcolepsy by logistic regression analysis. For each laboratory parameter, we used the most recent, maximum, and median values as predictors when fitting a generalized linear model with a binomial distribution. The models were adjusted for age at the time of measurement, sex, and the first ten PCs. To adjust for the confounding effect of BMI on laboratory measurements, we adjusted for BMI in a separate analysis.

### Medication prescriptions

Next, we added the medication data for every individual in the narcolepsy cohort extracted from purchase and reimbursement history. We selected the medications that are prescribed for the treatment of the cardiometabolic disorders. Similar to the epidemiological analysis of the cardiometabolic diseases diagnoses, medication purchase history was extracted both individually and as groups based on their anatomical therapeutic chemical (ATC) codes (https://atcddd.fhi.no/atc_ddd_index/). The groups included statins, insulins, GLP1 analogues, ADP inhibitors, beta blockers, calcium channel blockers, ACE inhibitors, and ARBs and ARB combinations. However, some medications remained uncategorized. From selected medications, statins and ezetimibe are associated with TG, LDL cholesterol and total cholesterol and insulins, GLP1 analogues, and metformin to blood glucose levels, indicative of metabolic diseases. ADP inhibitors, beta blockers, calcium channel blockers, ACE inhibitors and ARBs and their combinations, spironolactone and furosemide are all used to treat high blood pressure and its cardiovascular consequences. **Table 3** shows medication ATC codes and their prescription records across the narcolepsy cases and controls.

**Table 3.**
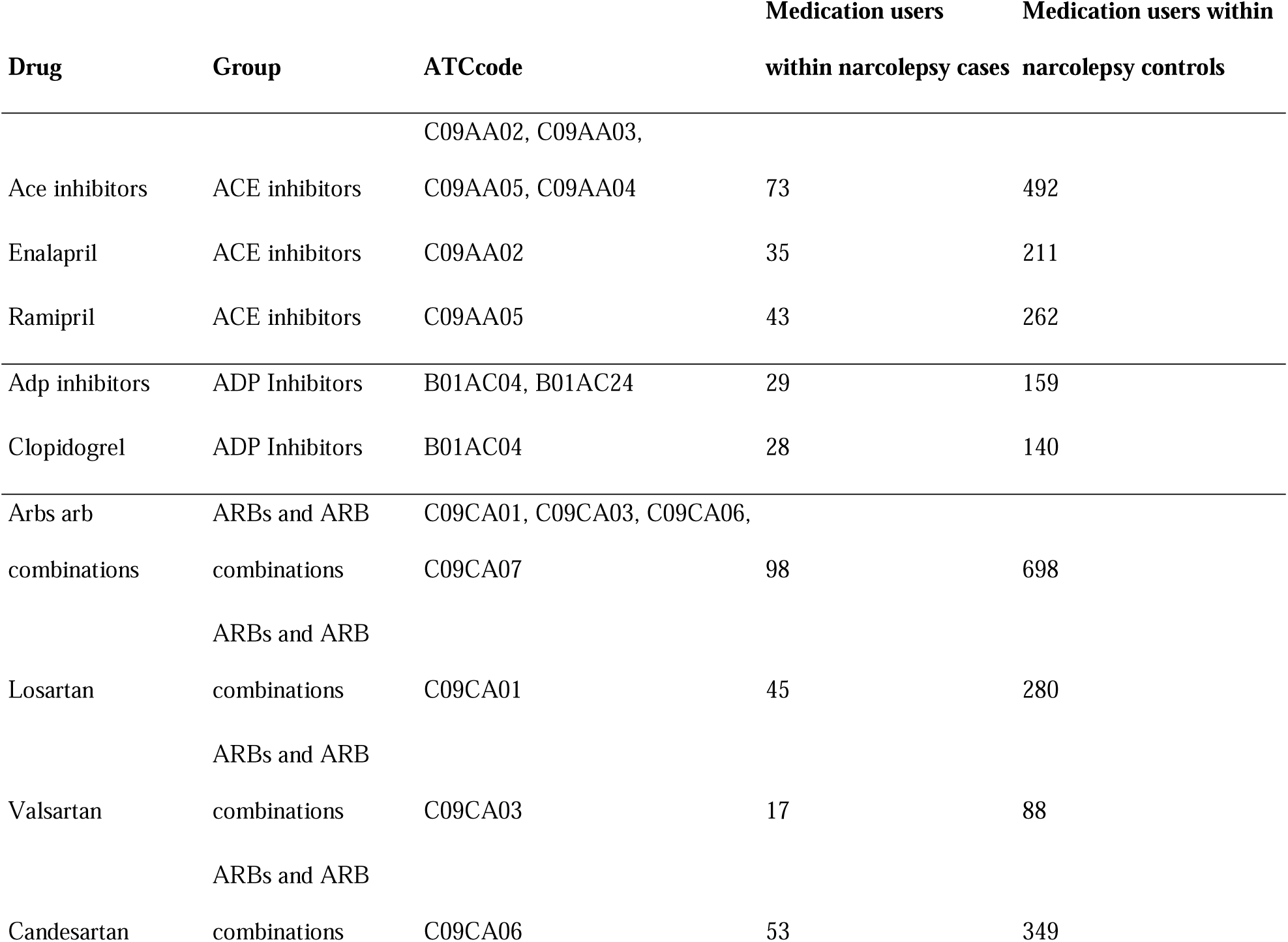
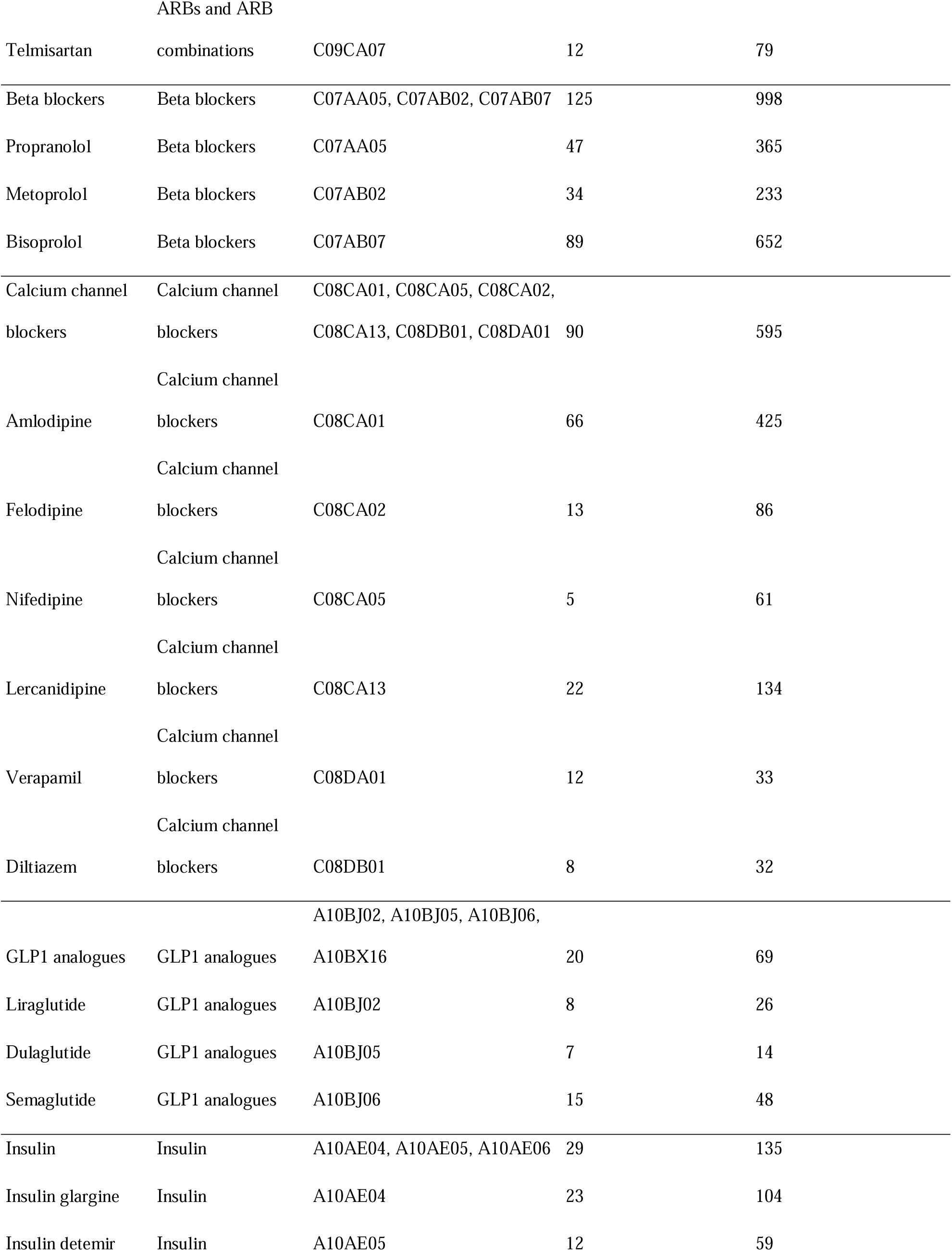
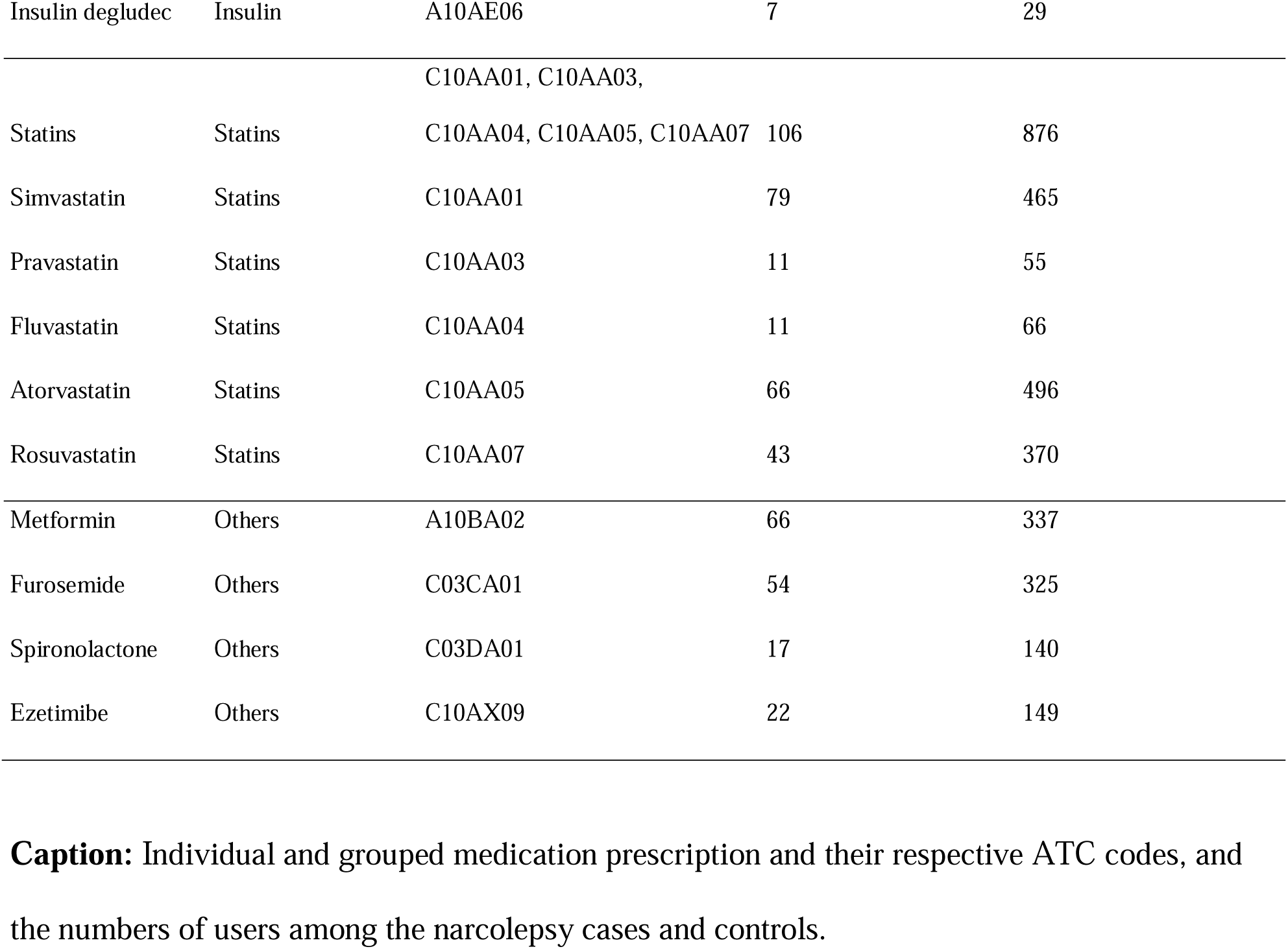
Medication prescription in narcolepsy cohort. Caption: Individual and grouped medication prescription and their respective ATC codes, and the numbers of users among the narcolepsy cases and controls.

### Association between narcolepsy and medication prescription

To investigate the association between narcolepsy and the selected medication prescriptions, we performed logistic regression by fitting a generalized linear model with a binomial distribution. Age, sex, and the first ten PCs were included as covariates. Similar to the previous sections, two sets of analyses, with and without adjusting for BMI, were performed.

## Results

### Narcolepsy is associated with cardiometabolic diseases

We examined the association between narcolepsy and cardiometabolic disorders in FinnGen using ICD-based definitions for most common cardiovascular and metabolic diseases (**Figure 2**). After multiple testing corrections using the false discovery rate (FDR) method, narcolepsy was associated with metabolic traits (and individually with obesity and T2D) and sudden death of unknown reasons. After additional adjustment for BMI, only associations with T2D, and the metabolic traits group remained significant. In addition, association between narcolepsy and stroke was significant in the BMI adjusted model. Results for BMI adjusted and unadjusted models for individual diagnoses and groups are presented in **Table S1**.

**Figure 2.**
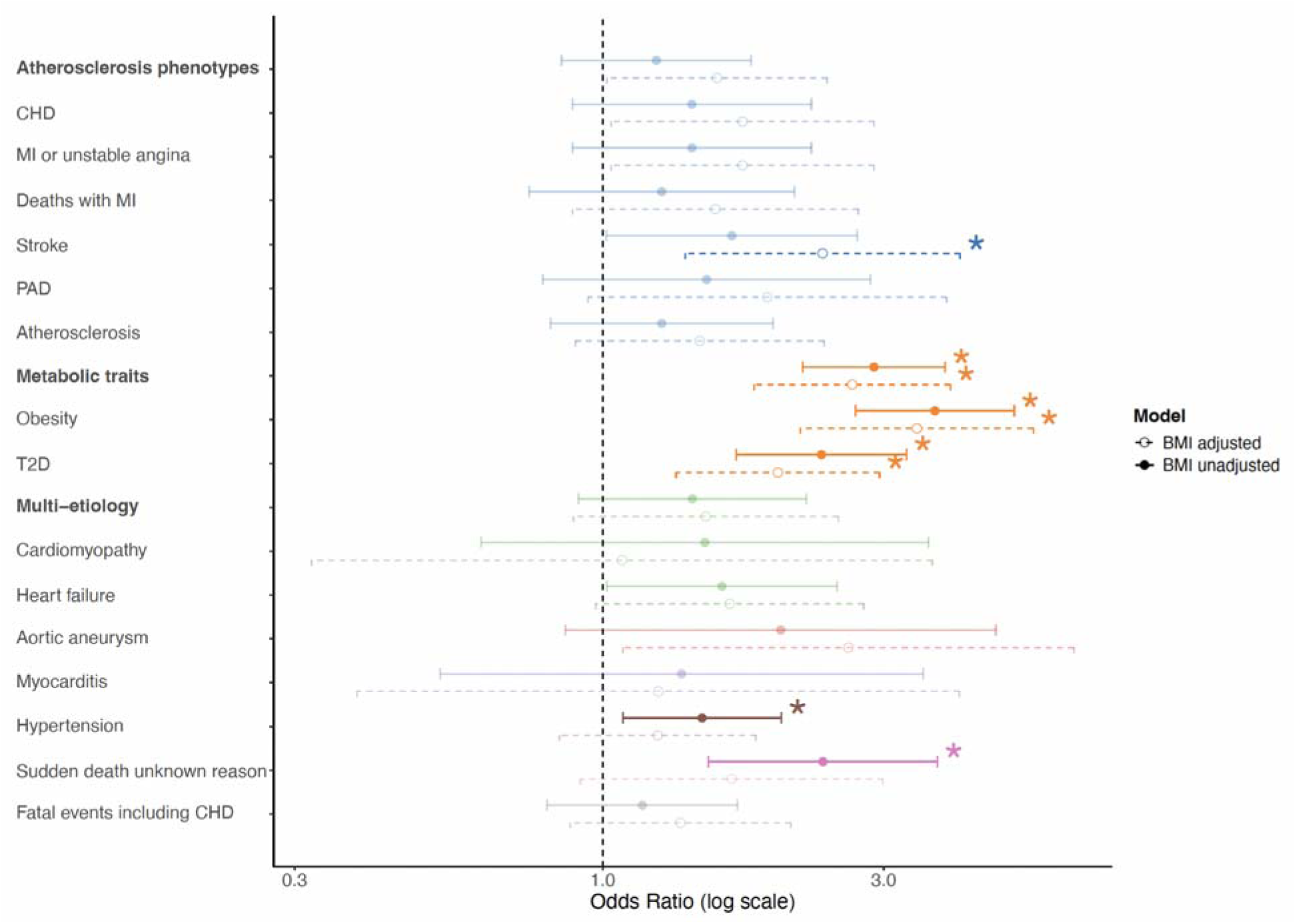
Epidemiological links between narcolepsy and cardiometabolic disorders. Forest plot displaying the odds ratios (log scale) for individual and grouped cardiometabolic diagnoses. Associations were assessed both with and without adjusting for BMI as a covariate. Phenotypes reaching statistical significance after correction for multiple testing are denoted by an asterisk (*).

### Narcolepsy is associated with higher glucose and lower HDL levels

Next, we studied the association between narcolepsy and laboratory values, indicative of cardiac and metabolic health. We included the last, maximum, and median laboratory measurement available in the Kanta dataset for each test (**Figure 3**).

**Figure 3.**
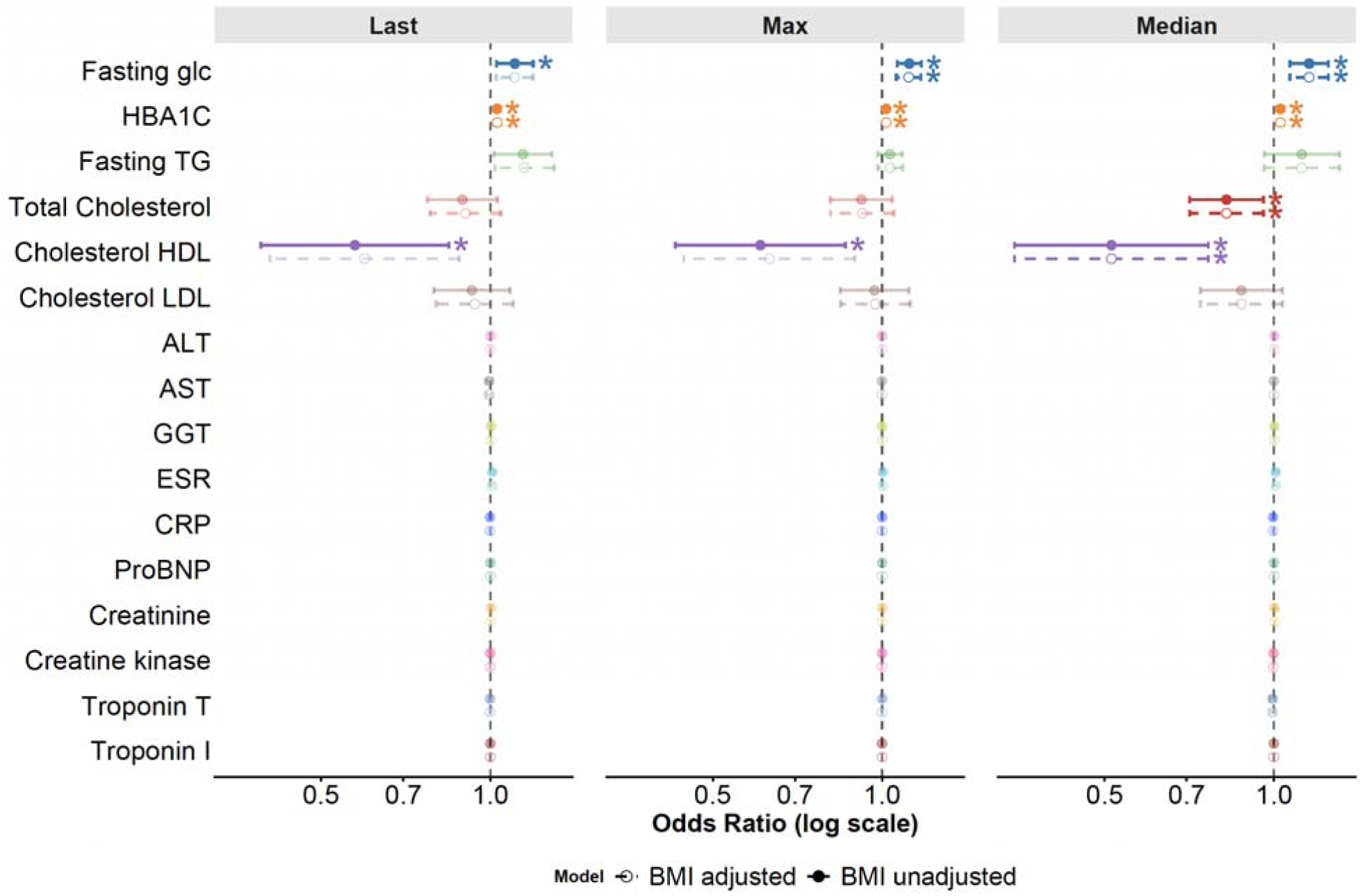
Association between laboratory measurements and narcolepsy diagnosis. Forest plots displaying the odds ratios (log scale) for last, maximum, and median values for each selected test. Results are shown with and without BMI as an additional covariate. The values that remain significant after multiple testing corrections are marked with an asterisk.

After FDR correction we observed higher fasting glucose levels within all measurement points with maximum measurement value to be statistically most significant. The association remained significant after additional adjustment with BMI. Similarly, significant association with HbA1c in all three timepoints, before and after adjusting for BMI was found, with the maximum measurement to be the most significant.

We also observed lower HDL cholesterol in narcolepsy vs. controls in all timepoints before BMI adjustment (strongest association with the median measurement value), remaining significant after additional adjustment for BMI. Negative association was also shown with total cholesterol in the median timepoint before and after adjusting with BMI.

Full statistical values for BMI adjusted and unadjusted models with last, maximum, and median measurements are presented in **Table S2**.

### Narcolepsy is associated with increased use of blood pressure and glucose lowering medications

Lastly, we leveraged medication prescriptions for selected cardiovascular and metabolic drugs. These were used as a proxy for cardiometabolic dysfunction, to study the association between narcolepsy diagnosis and medication prescriptions in FinnGen. Analysis was performed using individual medications and corresponding groups, with and without BMI as a covariate (**Figure 4 and Table S3**). We found significant associations with Simvastatin, insulin group (including insulin Glargine and insulin Detemir), metformin, and the GLP1 analogues (including Liraglutide, Dulaglutide and Semaglutide). Additionally, narcolepsy was positively associated with the prescription of ADP inhibitors (including Clopidogrel), Calcium channel blockers group (including Amlodipine and Verapamil), ACE inhibitors (including Ramipril and Enalapril), ARBs and ARB combinations (including Losartan) and Furosemide.

**Figure 4.**
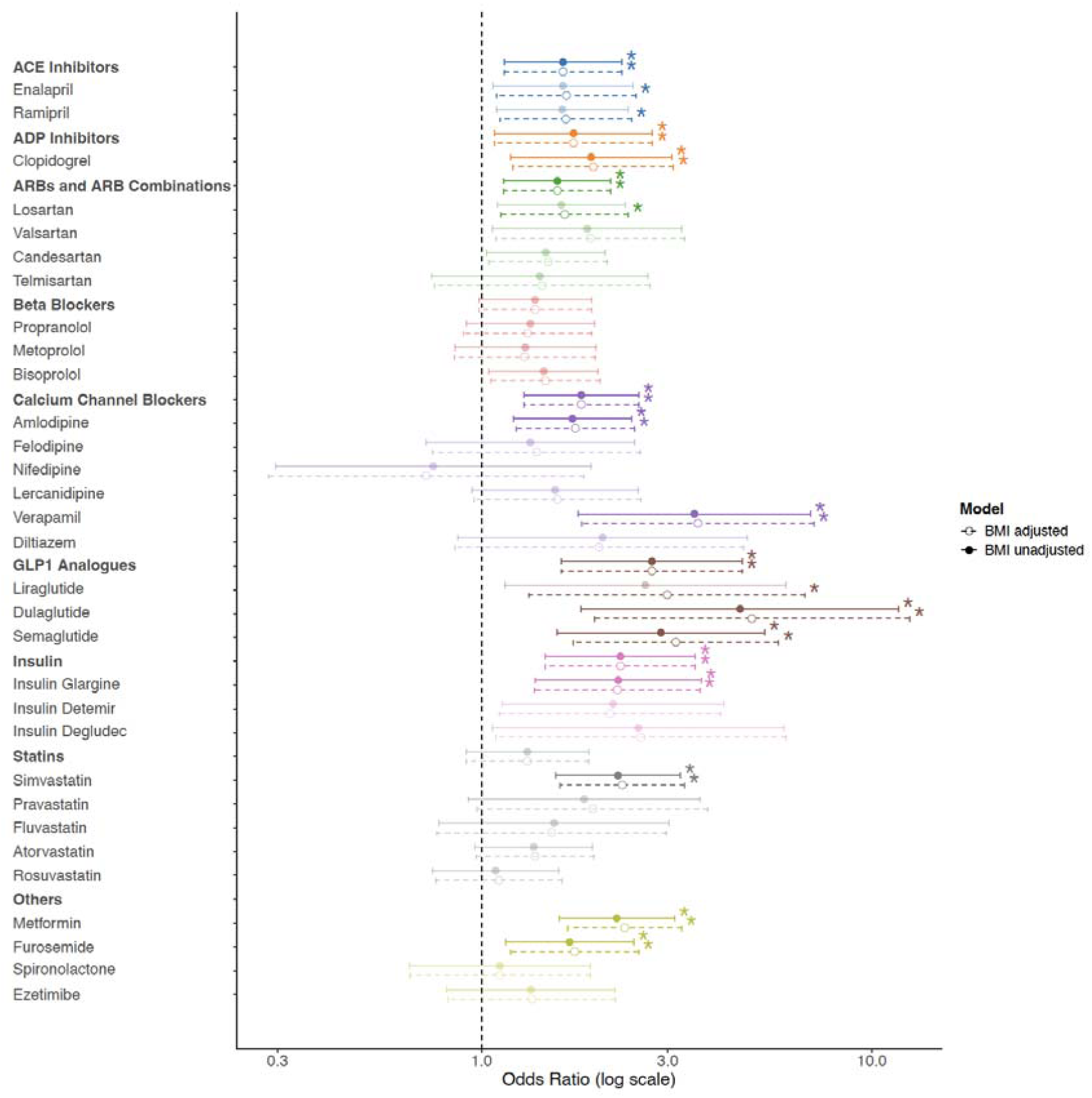
Epidemiological links between narcolepsy and cardiometabolic medication prescriptions. Forest plot displaying the odds ratios (log scale) for individual and grouped medications that are prescribed to treat cardiometabolic disorders. Associations were assessed both with and without adjusting for BMI as a covariate. Associations reaching statistical significance after correction for multiple testing are denoted by an asterisk.

## Discussion

NT1 is characterized by severe fatigue and sleep disruption and by low to zero levels of hypocretin^4^. In addition to its role in maintaining wakefulness, hypocretin has a crucial role in regulating BP, HR and energy balance^13,14^, thus potentially leading to an increased incidence of cardiometabolic comorbidities in narcolepsy.

In the present study, we leveraged FinnGen longitudinal EHR data, comprising ICD-based diagnoses, Kanta laboratory measurements, and medication prescription history from over 500,000 Finns, to address the cardiometabolic burden in narcolepsy.

First, we observed significant associations between narcolepsy and obesity, T2D, stroke, and sudden death of unknown reasons. The association with metabolic traits is consistent with previous literature. For example, a recent meta-analysis by Zhang et al. reported an obesity prevalence of approximately 30% among patients with narcolepsy, along with increased incidence rates of diabetes, hypertension, and dyslipidemia (10-20%)^29^. Several mechanisms have been proposed to explain the increased prevalence of obesity-related comorbidities in patients with narcolepsy, including disrupted energy homeostasis, altered food intake, impaired glucose tolerance, reduced insulin sensitivity, and chronic low-grade inflammation^30,31^. Our findings are in line with these earlier observations and expands the earlier findings by connecting narcolepsy directly to cardiovascular diseases at the level of clinical laboratory measurements, drug prescriptions and formal diagnoses. All of these showed a systematic cardiovascular and cardiometabolic burden in narcolepsy.

Interestingly, although hypocretin overexpression has been associated with increased food intake, it paradoxically protects against a high-fat diet-induced obesity rather than promoting excessive weight gain^32^. This protective effect is likely mediated through enhanced insulin and leptin sensitivity and improved glucose metabolism. In contrast, hypocretin deficiency, which is characterized in NT1, is associated with a reduced basal metabolic rate and altered eating behavior, potentially leading to excessive calorie intake and an increased risk of metabolic complications in affected patients^30^.

Although the increased incidence of obesity and related metabolic conditions may be partly explained by hypocretin deficiency^33^, lifestyle factors likely also contribute. Excessive daytime sleepiness and cataplexy may affect the ability to participate in physical activities^34,35^, potentially contributing to the development of obesity and metabolic dysfunction among the patients^34,36^.

Increased incidence of stroke within narcoleptic patients has been demonstrated before. Results from the Burden Of Narcolepsy Disease (BOND) study, a large-scale data analysis of the medical claims in United States, highlights an increased risk for stroke, MI, cardiac arrest, heart failure, with an increase in coronary revascularization procedures^34,37^. We speculate that some of the cases with diagnosis of sudden death of unknown reasons (ICD10: R96) in our cohort have also died of cardiovascular reasons.

In addition, we saw increased prevalence of atherosclerosis phenotypes (CHD, MI, PAD, atherosclerosis), heart failure, cardiomyopathy and aortic aneurysm within narcolepsy patients, however, these results did not pass the multiple testing correction.

Furthermore, narcolepsy was positively associated with several antihypertensive medications. The association of narcolepsy with higher use of these medications, likely reflects the higher prevalence of the underlying cardiometabolic conditions. However, despite the observed associations with several antihypertensive medications, the association with hypertension was only nominally significant. However, previous studies have reported an increased incidence of hypertension in patients with narcolepsy^41^. This discrepancy may reflect incomplete recording of the diagnosis, different treatment approaches, or the fact that medication use can capture individuals at an elevated cardiovascular risk even in the absence of a formal hypertension diagnosis.

Taken together, we observed a strong association between narcolepsy and cardiovascular and metabolic traits including obesity and T2D, as well as identified links with stroke and sudden death of unknown reasons. Laboratory measurements supported the link with compromised metabolic health. Lower HDL cholesterol in narcolepsy cases reflects a possible metabolic syndrome and T2D, associated with an increased risk of atherosclerosis, coronary heart disease, heart attack, and stroke. Finally, we observed increased purchases of the drugs targeting metabolic conditions, anti-hypertensives, and antiplatelet medications reinforcing associations with cardiometabolic health.

Our study is limited by the relatively small number of narcolepsy cases in the FinnGen cohort (285 diagnosed individuals), and the possible inclusion of both NT1 and NT2 patients that may dilute the observed associations. The sample size restricts the number of co-occurring cardiometabolic diagnoses and together with the clinical heterogeneity may reduce power to identify associations with statistical significance. It is noteworthy that we saw a strong association between cardiovascular parameters and narcolepsy even in this relatively small population. In addition, laboratory measurements in FinnGen are only available from 2014 onward, restricting our ability to compare values before and after narcolepsy diagnosis in all cases. Additionally, this limits us to study the measurements around 2011 peak in narcolepsy diagnosis in Finland, associated with H1N1 vaccination. Despite these limitations, the combination of clinical, laboratory, and prescription data highlights the importance to recognize cardiometabolic health as part of narcolepsy.

## Conclusion

To conclude, we show the association between narcolepsy and the most common cardiovascular disorders, including atherosclerosis phenotypes and metabolic traits including obesity and T2D. In addition, laboratory measurement patterns aligned with cardiovascular risk in patients compared to controls, showing a strong link between narcolepsy and the adverse lipid and glucose metabolism in narcolepsy. Finally, we observed an increased purchase of medications primarily prescribed to modulate the higher glucose levels in narcolepsy. These findings shed light on the cardiometabolic burden in narcolepsy and are beneficial to the medical sector to improve the treatment options and identify comorbid disorders.

## Acknowledgements

We want to acknowledge the participants and investigators of the FinnGen study. The FinnGen project is funded by two grants from Business Finland (HUS 4685/31/2016 and UH 4386/31/2016) and the following industry partners: AbbVie Inc., Alnylam Pharmaceuticals, Inc., AstraZeneca UK Ltd, Bayer AG, Biogen MA Inc., Boehringer Ingelheim International GmbH, Bristol Myers Squibb Inc. (and Celgene Corporation & Celgene International II Sàrl), Genentech Inc., GlaxoSmithKline Intellectual Property Development Ltd., Johnson&Johnson Innovative Medicine Inc., Maze Therapeutics Inc., Merck Sharp & Dohme LCC, Novartis AG, Pfizer Inc. and Sanofi US Services Inc. Following biobanks are acknowledged for delivering biobank samples to FinnGen: Auria Biobank (www.auria.fi/biopankki), THL Biobank (www.thl.fi/biobank), Helsinki Biobank (www.helsinginbiopankki.fi), Biobank Borealis of Northern Finland (https://www.ppshp.fi/Tutkimus-ja-opetus/Biopankki/Pages/Biobank-Borealis-briefly-in-English.aspx), Finnish Clinical Biobank Tampere (www.tays.fi/en-US/Research_and_development/Finnish_Clinical_Biobank_Tampere), Biobank of Eastern Finland (www.ita-suomenbiopankki.fi/en), Central Finland Biobank (www.ksshp.fi/fi-FI/Potilaalle/Biopankki), Finnish Red Cross Blood Service Biobank (www.veripalvelu.fi/verenluovutus/biopankkitoiminta), Terveystalo Biobank (www.terveystalo.com/fi/Yritystietoa/Terveystalo-Biopankki/Biopankki/) and Arctic Biobank (https://www.oulu.fi/en/university/faculties-and-units/faculty-medicine/northern-finland-birth-cohorts-and-arctic-biobank). All Finnish Biobanks are members of BBMRI.fi infrastructure (https://www.bbmri-eric.eu/national-nodes/finland/). Finnish Biobank Cooperative-FINBB (https://finbb.fi/) is the coordinator of BBMRI-ERIC operations in Finland. The Finnish biobank data can be accessed through the Fingenious® services (https://site.fingenious.fi/en/) managed by FINBB.

## FinnGen ethics statement

Study subjects in FinnGen provided informed consent for biobank research, based on the Finnish Biobank Act. Alternatively, separate research cohorts, collected prior the Finnish Biobank Act came into effect (in September 2013) and start of FinnGen (August 2017), were collected based on study-specific consents and later transferred to the Finnish biobanks after approval by Fimea (Finnish Medicines Agency), the National Supervisory Authority for Welfare and Health. Recruitment protocols followed the biobank protocols approved by Fimea. The Coordinating Ethics Committee of the Hospital District of Helsinki and Uusimaa (HUS) statement number for the FinnGen study is Nr HUS/990/2017. The FinnGen study is approved by Finnish Institute for Health and Welfare (permit numbers: THL/2031/6.02.00/2017, THL/1101/5.05.00/2017, THL/341/6.02.00/2018, THL/2222/6.02.00/2018, THL/283/6.02.00/2019,THL/1721/5.05.00/2019 and THL/1524/5.05.00/2020), Digital and population data service agency (permit numbers: VRK43431/2017-3, VRK/6909/2018-3, VRK/4415/2019-3), the Social Insurance Institution (permit numbers: KELA 58/522/2017, KELA 131/522/2018, KELA 70/522/2019, KELA 98/522/2019, KELA 134/522/2019, KELA 138/522/2019, KELA 2/522/2020, KELA 16/522/2020), Findata permit numbers THL/2364/14.02/2020, THL/4055/14.06.00/2020, THL/3433/14.06.00/2020, THL/4432/14.06/2020, THL/5189/14.06/2020, THL/5894/14.06.00/2020, THL/6619/14.06.00/2020, THL/209/14.06.00/2021, THL/688/14.06.00/2021, THL/1284/14.06.00/2021, THL/1965/14.06.00/2021, THL/5546/14.02.00/2020, THL/2658/14.06.00/2021, THL/4235/14.06.00/2021, Statistics Finland (permit numbers: TK-53-1041-17 and TK/143/07.03.00/2020 (earlier TK-53-90-20) TK/1735/07.03.00/2021, TK/3112/07.03.00/2021) and Finnish Registry for Kidney Diseases permission/extract from the meeting minutes on 4th July 2019.

The Biobank Access Decisions for FinnGen samples and data utilized in FinnGen Data Freeze 12 include: THL Biobank BB2017_55, BB2017_111, BB2018_19, BB_2018_34, BB_2018_67, BB2018_71, BB2019_7, BB2019_8, BB2019_26, BB2020_1, BB2021_65, Finnish Red Cross Blood Service Biobank 7.12.2017, Helsinki Biobank HUS/359/2017, HUS/248/2020, HUS/430/2021 28, 29, HUS/150/2022 12, 13, 14, 15, 16, 17, 18, 23, 58, 59, HUS/128/2023 18, Auria Biobank AB17-5154 and amendment #1 (August 17 2020) and amendments BB_2021-0140, BB_2021-0156 (August 26 2021, Feb 2 2022), BB_2021-0169, BB_2021-0179, BB_2021-0161, AB20-5926 and amendment #1 (April 23 2020) and it’s modifications (Sep 22 2021), BB_2022-0262, BB_2022-0256, Biobank Borealis of Northern Finland_2017_1013, 2021_5010, 2021_5010 Amendment, 2021_5018, 2021_5018 Amendment, 2021_5015, 2021_5015 Amendment, 2021_5015 Amendment_2, 2021_5023, 2021_5023 Amendment, 2021_5023 Amendment_2, 2021_5017, 2021_5017 Amendment, 2022_6001, 2022_6001 Amendment, 2022_6006 Amendment, 2022_6006 Amendment, 2022_6006 Amendment_2, BB22-0067, 2022_0262, 2022_0262 Amendment, Biobank of Eastern Finland 1186/2018 and amendment 22/2020, 53/2021, 13/2022, 14/2022, 15/2022, 27/2022, 28/2022, 29/2022, 33/2022, 35/2022, 36/2022, 37/2022, 39/2022, 7/2023, 32/2023, 33/2023, 34/2023, 35/2023, 36/2023, 37/2023, 38/2023, 39/2023, 40/2023, 41/2023, Finnish Clinical Biobank Tampere MH0004 and amendments (21.02.2020 & 06.10.2020), BB2021-0140 8/2021, 9/2021, 9/2022, 10/2022, 12/2022, 13/2022, 20/2022, 21/2022, 22/2022, 23/2022, 28/2022, 29/2022, 30/2022, 31/2022, 32/2022, 38/2022, 40/2022, 42/2022, 1/2023, Central Finland Biobank 1-2017, BB_2021-0161, BB_2021-0169, BB_2021-0179, BB_2021-0170, BB_2022-0256, BB_2022-0262, BB22-0067, Decision allowing to continue data processing until 31st Aug 2024 for projects: BB_2021-0179, BB22-0067,BB_2022-0262, BB_2021-0170, BB_2021-0164, BB_2021-0161, and BB_2021-0169, and Terveystalo Biobank STB 2018001 and amendment 25th Aug 2020, Finnish Hematological Registry and Clinical Biobank decision 18th June 2021, Arctic biobank P0844: ARC_2021_1001.

## Data availability

Data and code used in this study are available upon reasonable request. The FinnGen individual-level data may be accessed through applications to the Finnish Biobanks’FinnBB portal, Fingenious (www.finbb.fi). Summary data can be accessed through the FinnGen website https://www.finngen.fi/en/access_results.

